# Cardiac Function Consequences of the Persistence of Acute Myocarditis Somatostatin-PET Criteria Four Months Post-Acute Phase

**DOI:** 10.1101/2024.09.19.24314017

**Authors:** Thomas Larive, Caroline Boursier, Marine Claudin, Jeanne Varlot, Laura Filippetti, Olivier Huttin, Véronique Roch, Laetitia Imbert, Matthieu Doyen, Antoine Fraix, Damien Mandry, Elodie Chevalier, Pierre-Yves Marie

**Affiliations:** Université de Lorraine, Department of Nuclear Medicine and Nancyclotep Imaging Platform, CHRU Nancy, F-54000, Nancy, France; Université de Lorraine, IADI, INSERM U1254, F-54000, Nancy, France; Department of Cardiology, CHRU Nancy, F-54000, Nancy, France; Université de Lorraine, INSERM, UMR 1116, 54000, Nancy, France; Université de Lorraine, Department of Radiology, Brabois, CHRU Nancy, F-54000, Nancy, France; Université de Lorraine, CHRU-Nancy, INSERM, CIC 1433, Nancy, France

**Keywords:** [^68^Ga]Ga-DOTA-TOC, PET, myocardial inflammation, somatostatin receptor, acute myocarditis

## Abstract

**Background:** Somatostatin-Positron Emission Tomography (PET) imaging of inflammatory cells is an effective approach for detecting Acute Myocarditis (AM), based on the Myocardial Uptake Volume (MUV) criteria of > 18 cm^3^. The current study further characterizes patients for whom this criterion persists at the 4-month Follow-Up (FU) from apparently uncomplicated AM.

**Methods:** Twenty-seven patients [median age 26.5, inter-quartile range: 21.9-31.9 years], underwent Cardiac Magnetic Resonance (CMR) and [^68^Ga]Ga-DOTA-TOC PET at the acute phase and 4.5 [4.2-5.0] months later. Patients with > 18 cm^3^ MUV (FU.PET+) at the 4-month follow-up were compared to FU.PET-patients.

**Results:** At 4 months, inflammation by CMR was only identified in two patients but in 11 patients by PET (FU.PET+ group), with 5 of these 11 patients exhibiting a baseline-to-4-months expansion in MUV. Ejection Fractions (EFs) at 4 months were generally lower in FU.PET+ than FU.PET-patients (Left Ventricular (LV).EF, 52.9 [48.6; 55.0] % vs. 56.0 [54.3; 57.8] %, p=0.001). In addition, the 5 FU.PET+ patients with expansion of the MUV had a worse LV.EF evolution (4-month follow-up minus baseline difference in LV.EF: (−5.0) [(−12.9)-(−1.0)] %) vs. + 3.1 [0.94-8.0] % for the other patients, p=0.004) and the highest plasma high-sensitivity troponin-Ic at 4-months (13.0 [7.5-22.00] ng/l vs. 3.0 [2.0-9.5] ng/l, p=0.045) suggestive of a more persistent active disease. In contrast, the evolution profile of the 6-remaining FU.PET+ patients was consistent with a longer recovery time as a consequence of a more severe initial insult (i.e. with lower LV.EF and higher MUV at baseline vs. the other patients, both p < 0.05).

**Conclusions:** Myocardial inflammation is detected by somatostatin-PET at 4 months from an apparently uncomplicated AM in as many as 41% of patients. It is associated with a poorer recovery of cardiac function, consistent with more persistent active disease or a more severe initial insult.

**Clinical Perspective:** Although generally of mild severity, myocarditis may potentially involve dreaded complications such as heart failure, arrhythmia, and sudden cardiac death. We recently showed that the Somatostatin-positron emission tomography (PET) determination of inflammatory cell volume constitutes an alternative to cardiac magnetic resonance (CMR) imaging for detecting myocarditis at the acute phase. The present study shows that PET criteria of myocardial inflammation persist at four months from apparently uncomplicated myocarditis at a much higher rate than CMR (41% vs. 7% of patients). Moreover, this persistence is associated with a poorer recovery of cardiac function and is in line with two different evolution profiles—i.e., a more persistent active disease or a more severe initial insult. These new findings have the potential to improve monitoring and treatment adaptations for myocarditis patients.

**Graphical abstract:** 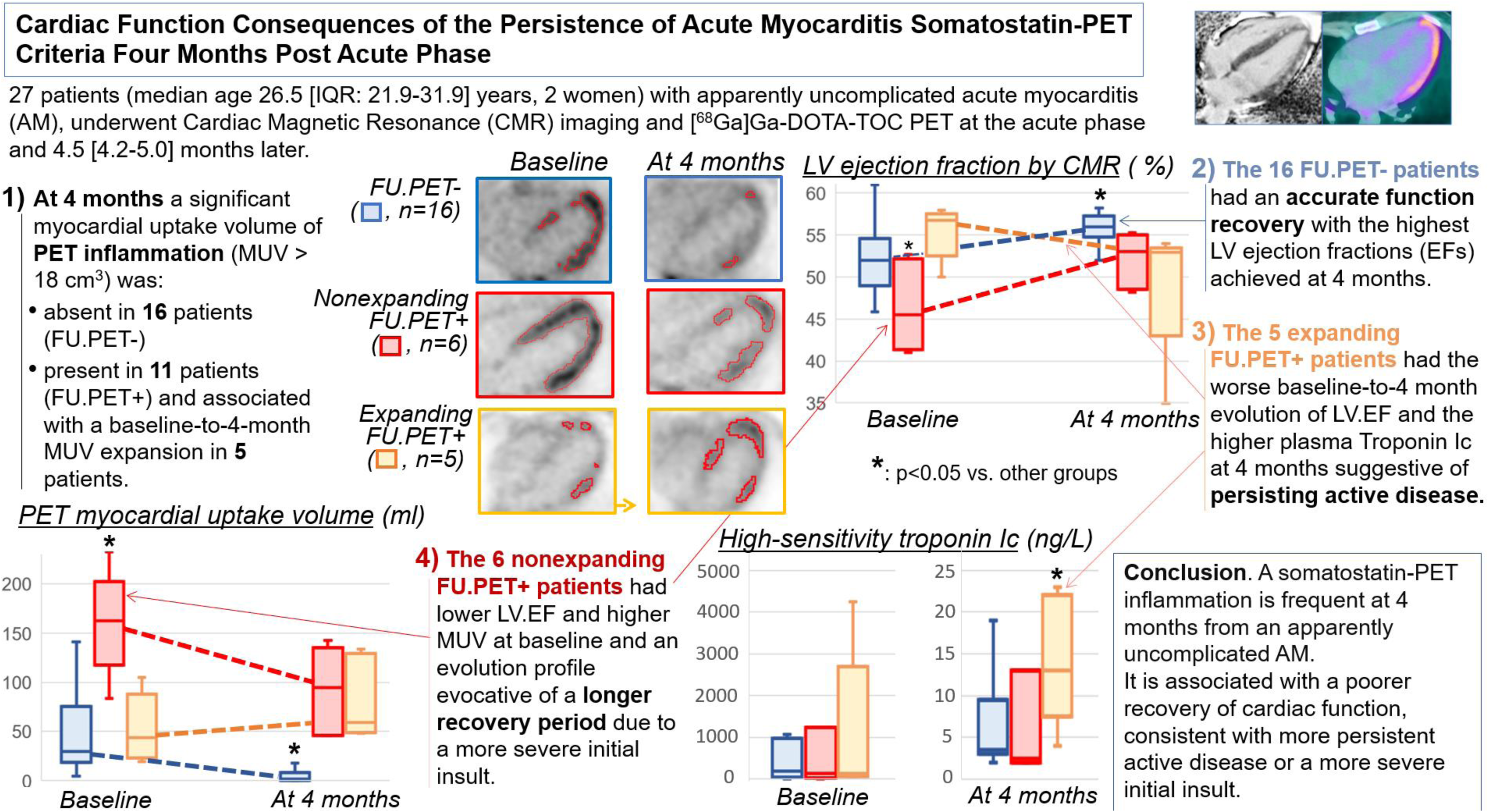

## Introduction

Myocarditis from infectious or noninfectious causes is generally of mild severity but may potentially involve dreaded complications such as heart failure, arrhythmia, and sudden cardiac death. Early detection of Acute Myocarditis (AM) is essential and has been facilitated by the advent of Cardiac Magnetic Resonance (CMR) imaging and image analysis based on the modified Lake-Louise criteria. However, this CMR methodology has low sensitivity for detecting subacute or chronic myocarditis, particularly in patients with cardiomyopathy-like or arrhythmia presentations (*1*)

Positron Emission Tomography (PET) imaging of somatostatin receptors was initially developed for assessing neuroendocrine tumors. However, somatostatin-PET also detects inflammatory cells overexpressing somatostatin receptors, such as activated macrophages and lymphocytes, which are the primary cell subsets implicated in myocarditis (*2*,*3*). We recently showed that somatostatin-PET could constitute an alternative to CMR for identifying myocarditis at the acute phase when the volume and intensity of myocardial somatostatin-PET uptake are quantified relative to blood activity (*4,5*). This relative quantification correlates better with somatostatin receptor density than standardized uptake values (SUV) (*6*).

Perhaps a more surprising preliminary observation was that PET signs of myocardial inflammation persisted at 4 months from the acute phase in many of our patients, at a time when inflammation was no longer identified by CMR and when the healing phase is generally considered to be completed (*5*). This raised the question whether somatostatin-PET could unmask chronic forms of myocarditis that would be significant enough to affect cardiac function.

This dual somatostatin-PET/CMR study therefore aimed to further characterize patients that satisfied the AM PET criteria at 4 months from apparently uncomplicated AM, specifically in terms of consequences on cardiac function.

## Methods

This prospective study initially identified 32 patients, hospitalized for Acute Myocarditis (AM) that fulfilled the 2018 acute myocarditis Lake Louise CMR criteria, with significant increases in plasma troponin-Ic and no evident myocardial disease other than myocarditis.

Patients were referred for a [^68^Ga]Ga-DOTA-TOC PET ≤ 72 hours after the initial CMR, and 4 months later, on the day of the additional CMR. A venous blood sample was collected before each PET investigation to determine C Reactive Protein (CRP) and high-sensitivity troponin Ic levels.

The study protocol was approved by the ethics committee (“Comité de Protection des Personnes - Ouest IV”) and released on the clinical.trial.gov site under identifier NCT03347760. All participants signed an informed consent to participate.

### [^68^Ga]Ga-DOTA-TOC PET/CT

The somatostatin-PET recording and analysis methods are detailed elsewhere (*4,5*). To summarize, 1 hour post injection of 2 MBq/kg of [^68^Ga]Ga-DOTA-TOC, the CT was recorded first and followed by a single bed position 15 min cardiac PET acquired on a digital hybrid PET/CT system (Vereos, Philips, Cleveland, Ohio), with the patient preferentially set in the prone position. PET images were reconstructed as follows: 2 mm voxels, OSEM algorithm with 3 iterations and 3 subsets, a deconvolution of the point spread function and further corrections for scatter, random coincidences and attenuation.

Areas of myocardial [^68^Ga]Ga-DOTA-TOC uptake were assessed visually on a 0-to-≤3 scale and considered significant if they (i) were associated with a myocardial/blood Standardized Uptake Value (SUV) max ratio of > 1.5, with blood activity determined from a spherical, 2 cm diameter, volume of interest positioned at the center of the right atrium and (ii) were located at more than 1 cm from liver activity (*4*). PET Myocardial Uptake Volumes (MUVs) were thereafter quantified based on isocontours and by applying the above visual analysis criteria (i.e. myocardial/blood SUVmax ratio > 1.5 and > 1 cm from liver activity).

### CMR investigations

CMR images were recorded on a 1.5T Avento or a 3T Prisma system (Siemens, Erlangen, Germany) during the acute phase and at follow-up, as previously described by our team (*7*).

Briefly, LV and Right Ventricular (RV) functions as well as LV mass were measured from contiguous short-axis slices recorded with a compressed sensing SSFP sequence using AI-based software (Arterys, Tempus AI Inc., USA). LV mass and ventricular volumes were indexed to body surface area.

Longitudinal (T1) and transversal (T2) relaxation maps were recorded from short-axis slices with precontrast - Modified Look-Locker Inversion Recovery (MOLLI, acquisition scheme 5(3)3) and 2D TurboFlash sequences, respectively (*7*). Myocardial T1 and T2 were determined from Regions Of Interest (ROIs) outlined on a septal mid-ventricular area, as well as LV areas with Late Gadolinium Enhancement (LGE). Since the 1.5 and 3T scanners have different normal T1 and T2 ranges, we normalized and expressed these parameters as percentages of previously defined mean normal values (i.e. on the 3T scanner, 1256 and 38.5 ms, for T1 and T2, respectively (*7,8*); on the 1.5T scanner, 1025 and 45.2 ms, for T1 and T2, respectively (*9,10*)). The upper normal T1 and T2 thresholds corresponded to the upper 95% confidence interval limits from these normal values.

Contiguous short- and long-axis LGE images, covering the entire LV volume, were recorded with a fast-multi-slice phase-sensitive inversion recovery sequence (*7*), 10 to 15 min post injection of 0.1 mmol.kg^−1^ body weight of Dotarem®, (GUERBET, France). Segments with LGE were identified visually based on a 17-segment division of the LV (*11*).

### Statistical analysis

Qualitative variables are expressed as numbers and percentages, and quantitative variables as medians with interquartile ranges. Wilcoxon and Mann-Whitney tests were used to compare paired and unpaired quantitative variables, respectively. P values < 0.05 were considered to reflect significant differences.

## Results

### Baseline data

Out of the 32 patients initially identified, 1 patient was excluded because of image archiving issues and 4 patients opted out of the 4-month PET follow-up. A total of 27 patients (median age 26.5 [21.9-31.9] years, 2 women) were therefore considered in the analysis. Previous medical history included coronary angioplasty (n=1) and acute myocarditis (n=1) and 12 (44%) patients had cardiovascular risk factors at the time of acute phase disease, i.e. 6 obese patients (body mass index > 30 kg/m^2^), 5 active smokers, and 1 patient with medically treated hypertension.

All patients presented with elevated plasma troponin-Ic levels during the acute phase (median peak: 12.0 [4.8-20] ng/mL). All initial CMR scans performed at a median of 2 [1-3] days from peak troponin, exhibited characteristic sub-epicardial LGEs together with increased myocardial T2s, thereby fulfilling the CMR diagnostic criteria of acute myocarditis (*12*). Nine patients (32%) had an LV ejection fraction of < 50%.

The somatostatin-PET, performed at a median of 4 [3-6] days post peak troponin, identified the presence of a volumetric AM criterion (i.e. MUV > 18 cm^3^) in 85% (23/27) of patients.

### The 4-month evolution

At the 4-month follow-up, only 2 patients had elevated troponin-Ic levels (75 and 63 pg/ml compared to an upper normal threshold of 40 pg/ml) and 3 patients had a C-reactive protein (CRP) level above the normal range (10, 14 and 22 mg/L compared to an upper normal threshold of 9 mg/l). CMR showed the persistence of LGE in all patients, with only 3 (11%) patients presenting an LV.EF of < 50% and 2 (7%) patients with an increased myocardial T2. As detailed in Table 1, there were significant improvements in both the LV and Right Ventricular (RV) EFs, in the LV mass and LV end-systolic volumes together with marked decreases in the number of LGE segments and in myocardial T1 and T2.

**Table 1:** Main continuous variables and PET data obtained from the overall population (expressed as medians [with interquartile ranges]) and P values for paired comparisons between the acute phase vs. the 4-month follow-up.

|  | **Acute phase**<br>(n=27) | **4-month follow-up**<br>(n=27) | P value |
| --- | --- | --- | --- |
| Troponin Ic (ng/l) * | 125 [39-1074] | 4 [3-13] | < 0.001 |
| C-reactive protein (mg/l) * | 5.0 [0.0-13.8] | 0.0 [0.0-0.0] | 0.009 |
| **CMR data** |  |  |  |
| Interval from troponin peak (in days) | 2.0 [1.0-3.0] | 139 [132-153] | < 0.001 |
| Heart rate (bpm) | 60 [56-75] | 58 [50-71] | 0.081 |
| Indexed stroke volume (mL.m<sup>-2</sup>) | 40.9 [37.9-47.0] | 44.4 [39.6-47.3] | 0.098 |
| Indexed LV.EDV (mL.m<sup>-2</sup>) | 82.8 [73.1-89.9] | 78.1 [69.3-83.4] | 0.064 |
| Indexed LV.ESV (mL.m<sup>-2</sup>) | 38.0 [34.5-45.3] | 34.7 [30.9-39.3] | 0.009 |
| LV.EF (%) | 52.0 [49.0-56.0] | 55.0 [52.2-57.0] | 0.038 |
| Indexed LV mass (g.m<sup>-2</sup>) | 56.4 [51.9-63.5] | 50.3 [46.3-58.0] | 0.001 |
| Indexed RV.EDV (mL.m<sup>-2</sup>) | 84.4 [69.0-93.5] | 81.9 [74.9-94.0] | 0.638 |
| Indexed RV.ESV (mL.m<sup>-2</sup>) | 43.2 [32.5-50.2] | 37.1 [33.5-47.1] | 0.144 |
| RV.EF (%) | 49.7 [45.0-51.9] | 51.8 [47.9-56.2] | 0.015 |
| T1 (%) | 115 [108-121] | 97 [95-99] | < 0.001 |
| T2 (%) | 135 [125-146] | 104 [99-117] | < 0.001 |
| Number of LGE segments | 5 [3-6] | 3 [2-4] | < 0.001 |
| **PET data** |  |  |  |
| Interval from troponin peak (in days) | 4.0 [3.0-6.0] | 139 [132-153] | < 0.001 |
| LV/blood SUV max ratio | 2.72 [2.31-2.99] | 1.92 [1.67-2.24] | < 0.001 |
| MUV (mL) | 47.3 [21.9-31.9] | 9.0 [0.8-59.5] | 0.004 |

EDV: end-diastolic volume, ESV: end-systolic volume, LV: left ventricle, MUV: myocardial uptake volume; RV: right ventricle, SUV: standardized uptake value, T1: myocardial longitudinal relaxation time, T2: myocardial T2 relaxation time.
\*: determined on the day of the PET investigations.

Significant, albeit less marked, decreases were also identified for the two somatostatin-PET variables: MUV and myocardial-to-blood SUVmax ratio (Table 1). Nevertheless, 11 patients (41%) satisfied a persisting acute myocarditis PET criterion at the 4-month follow-up (MUV > 18 cm^3^ (*4*), FU.PET+ group), whereas the remaining 16 patients did not (FU.PET-group).

### Characterization of the FU.PET+ patients

As detailed in Table 2 and Figure 1, significant 4-month follow-up improvements in EFs as well as LV and RV end-systolic volumes were observed in the FU.PET-group (all p<0.05) but not in the FU.PET+ group. Consequently, lower LV.EF (52.9 [48.6-55.0] % vs. 56.0 [54.3-57.8] %, p=0.001) and RV.EF (49.6 [45.9-51.8] % vs. 54.8 [50.6-57.1] %, p=0.023) follow-up levels were achieved in the FU.PET+ group compared to the FU.PET-patients.

**Figure 1:**
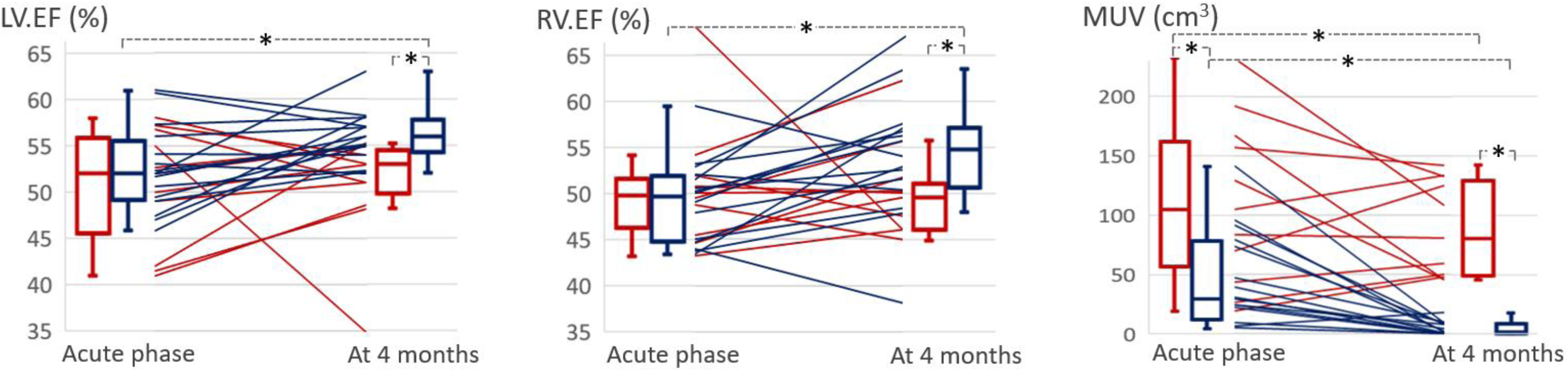
Box plots and individual evolution of left and right ventricular ejection fractions (LV.EF and RV.EF, respectively) and myocardial PET uptake volumes (MUV) between the acute phase and the 4-month follow-up for patients fulfilling the PET criteria of myocardial inflammation at 4 months (FU.PET+ group, lines and boxes in red) and patients with no persisting AM PET criteria (FU.PET-group, lines and boxes in blue). *: p < 0.05 for paired and unpaired comparisons.

**Table 2:** Main continuous variables and PET data obtained from patients that fulfill the AM PET criteria at the 4-month follow-up (FU.PET+ group) and patients with no persisting AM PET criteria (FU.PET-group) expressed as medians [with interquartile ranges]) and P values for paired comparisons between the acute phase vs. the 4-month follow-up.

|  | Acute phase |  |  | 4-month follow-up |  |  |
| --- | --- | --- | --- | --- | --- | --- |
|  | FU.PET +<br>(n=11) | FU.PET –<br>(n=16) | P value | FU.PET +<br>(n=11) | FU.PET –<br>(n=16) | P value |
| Age (years) | 31.4 [25.0-38.2] | 24.4 [20.7-31.2] | 0.080 | ----- | ----- | ----- |
| Body mass index (kg.m<sup>2</sup>) | 25.7 [24.8-31.2] | 24.8 [22.9-29.1] | 0.451 | ----- | ----- | ----- |
| High-sensitivity troponin Ic (ng/l) | 125 [44-1156] | 184 [29-1039] | 0.368 | 7 [2-21] * | 3 [3-11] * | 0.716 |
| C-reactive protein (mg/l) | 0.0 [0.0-6.8] | 10.9 [0.0-24.3] | 0.064 | 0.0 [0.0-0.0] | 0.0 [0.0-3.8] * | 0.451 |
| **CMR data** |  |  |  |  |  |  |
| Heart rate (bpm) | 59 [54-76] | 62 [56-69] | 0.790 | 57 [50-79] | 58 [50-67] | 0.878 |
| Indexed stroke volume (mL.m<sup>-2</sup>) | 39.7 [36.1-46.3] | 42.6 [38.2-47.7] | 0.451 | 41.7 [37.2-47.1] | 45.1 [40.6-47.4] | 0.318 |
| Indexed LV.EDV (mL.m<sup>-2</sup>) | 83.6 [73.3-93.6] | 82.0 [71.6-88.8] | 0.577 | 81.9 [70.3-95.8] | 76.7 [69.2-82.2] | 0.212 |
| Indexed LV.ESV (mL.m<sup>-2</sup>) | 37.2 [34.6-49.7] | 38.5 [33.2-42.9] | 0.610 | 38.8 [33.2-49.2] | 33.3 [30.3-36.7] * | 0.044 |
| LV.EF (%) | 52.0 [42.0-56.7] | 52.0 [49.0-55.5] | 0.680 | 52.9 [48.6-55.0] | 56.0 [54.3-57.8] * | 0.001 |
| Indexed LV mass (g.m<sup>-2</sup>) | 58.6 [54.3-72.0] | 54.9 [50.1-63.3] | 0.251 | 50.6 [48.2-61.3] * | 50.1 [44.3-56.0] * | 0.294 |
| Indexed RV.EDV (mL.m<sup>-2</sup>) | 81.8 [64.7-95.3] | 86.5 [72.0-92.8] | 0.660 | 88.1 [74.9-95.2] | 79.2 [73.3-85.2] | 0.178 |
| Indexed RV.ESV (mL.m<sup>-2</sup>) | 42.7 [31.5-50.2] | 43.9 [33.5-50.8] | 0.660 | 46.3 [36.2-51.5] | 36.2 [32.7-41.3] * | 0.050 |
| RV.EF (%) | 49.7 [45.2-52.4] | 49.7 [44.8-51.9] | 0.816 | 49.6 [45.9-51.8] | 54.8 [50.6-57.1] * | 0.023 |
| T1 (%) | 115 [105-122] | 115 [108-120] | 0.827 | 97 [95-102] * | 96 [95-98] * | 0.394 |
| T2 (%) | 133 [124-144] | 140 [128-148] | 0.421 | 106 [99-118] * | 103 [99-111] * | 0.451 |
| Number of LGE segments | 4 [3-7] | 5 [3-6] | 0.790 | 3 [2-4] * | 3 [2-4] * | 0.544 |
| **PET data** |  |  |  |  |  |  |
| Interval from troponin peak (in days) | 4.5 [3.0-5.8] | 4.0 [3.0-6.0] | 0.753 | 137 [132-146] | 152 [132-155] | 0.272 |
| LV/blood SUV max ratio | 2.96 [2.36-3.64] | 2.50 [2.26-2.92] | 0.099 | 2.25 [2.23-2.46] * | 1.73 [1.47-1.92] * | < 0.001 |
| MUV (mL) | 105.2 [43.4-163.3] | 29.9 [12.5-78.2] | 0.011 | 80.4 [47.8-133.0] | 1.7 [0.1-8.5] * | < 0.001 |

\*: p < 0.05 for paired comparisons between the acute phase and the 4-month follow-up. Abbreviations are the same as in Table 1

The analysis of individual variations in the graph on the right of Figure 1, shows that MUV was still expanding between baseline and the 4-month follow-up in 5 FU.PET+ patients (difference between 4 months and baseline: + 28.4 [19.9-41.8] cm^3^). As detailed in Figure 2, this expanding FU.PET+ subgroup was characterized by an even worse evolution of LV.EF compared to the other patients (4 months minus baseline difference in LV.EF: (−5.0) [(−12.9)-(−1.0)] %) vs. + 3.1 [0.94-8.0] % for the other groups, p=0.004). The evolution profile was very different in the 6 other and nonexpanding FU.PET+ patients with their LV.EF and MUV being more abnormal at baseline than in the other patients but clearly improved at 4 months (Figure 2 and graphical abstract).

**Figure 2:**
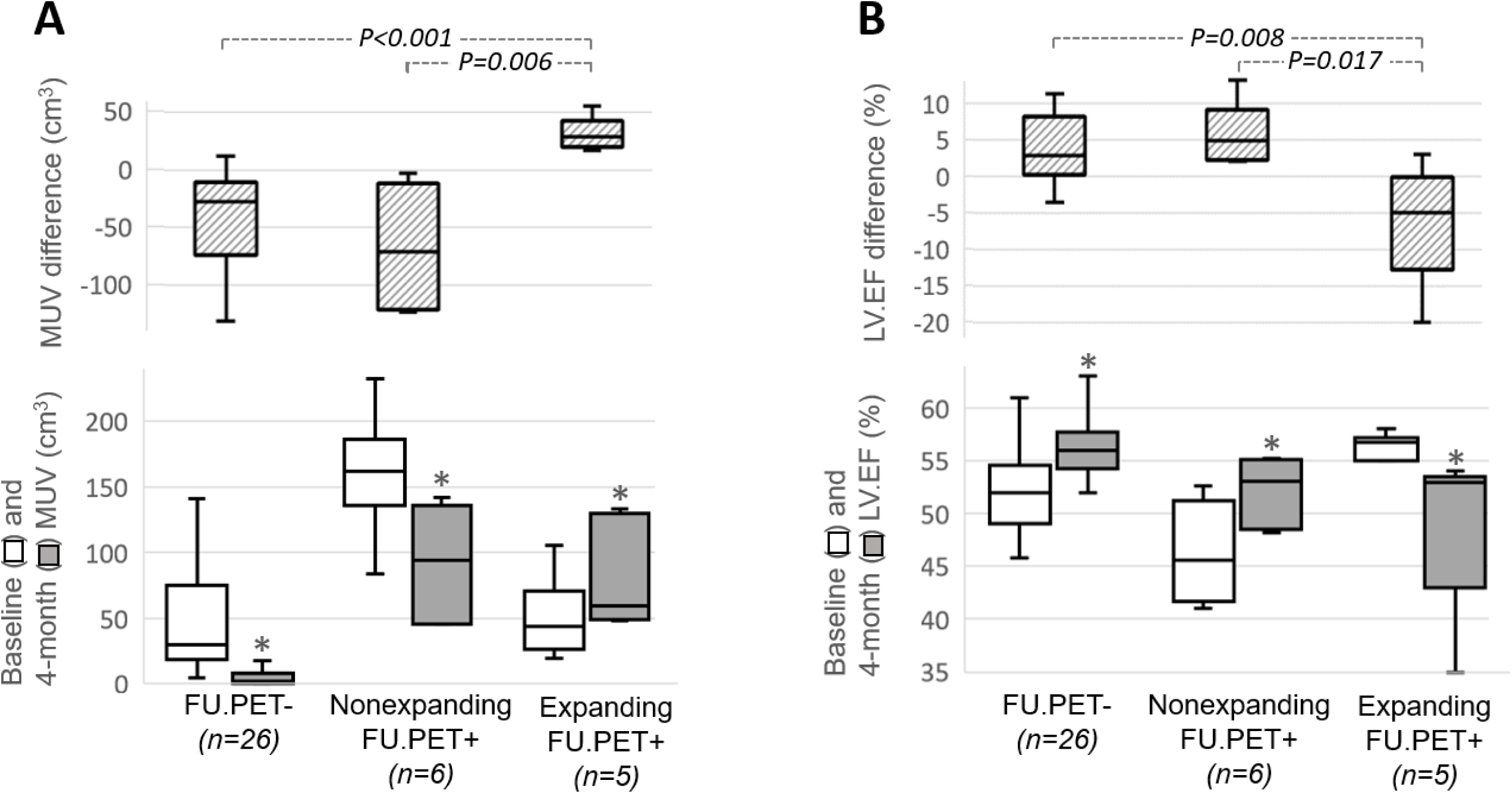
Box plots of (A) myocardial uptake volumes (MUV) and (B) left ventricular ejection fractions (LV.EF) between baseline and 4-month follow-up, as well as their 4-month-minus-baseline differences, for patients fulfilling the PET criteria of myocardial inflammation at 4 months (FU.PET+) and patients with no persisting AM PET criteria (FU.PET-). The FU.PET+ group was further dichotomized into subgroups with increased MUVs at 4 months compared to baseline (Expanding FU.PET+) and FU.PET+ patients with MUVs that did not increase from baseline (Nonexpanding FU.PET+). *: p < 0.05 for paired comparisons between baseline and 4 months.

**Figure 3:**
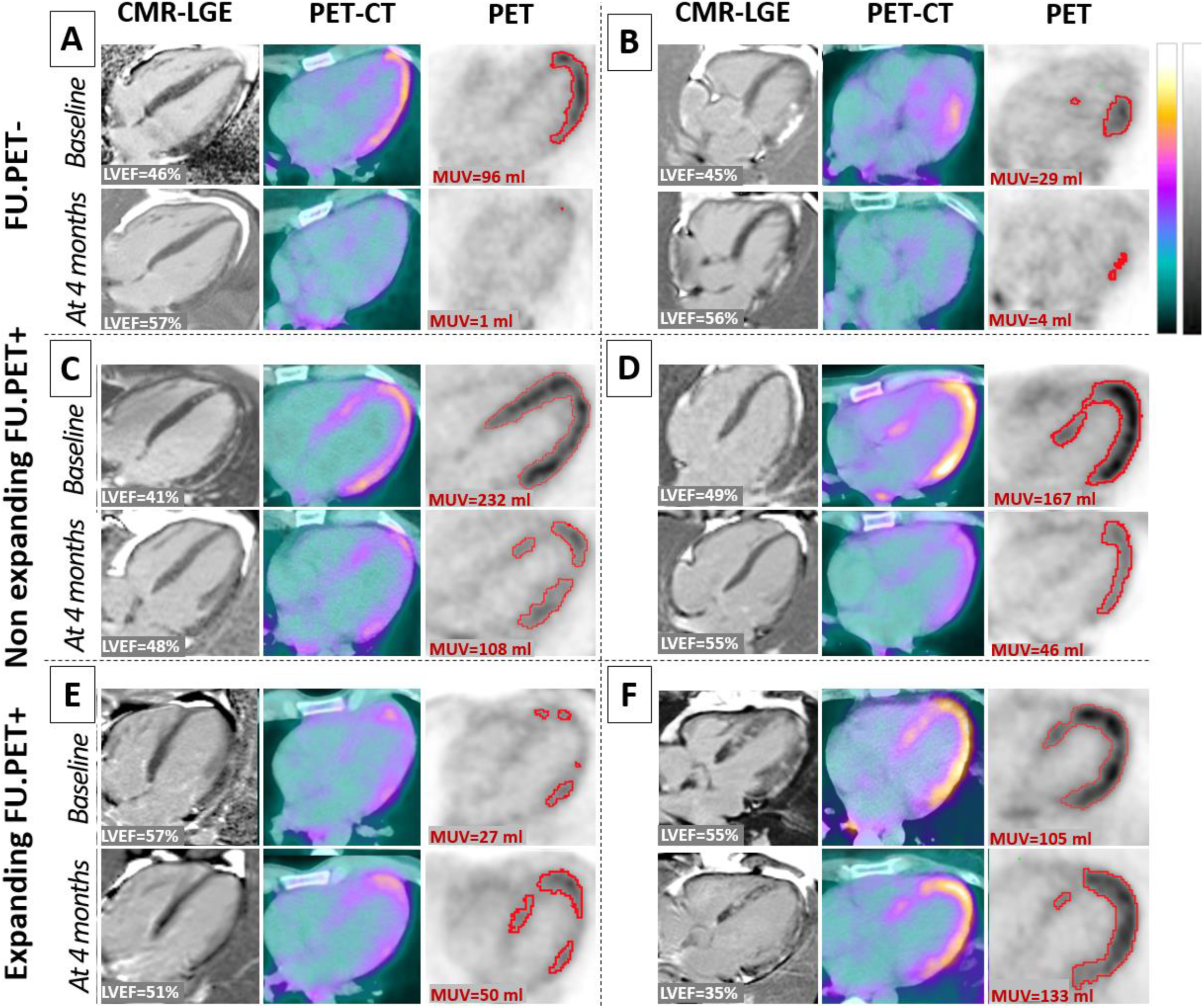
Representative horizontal long-axis slices recorded at baseline and at the 4-month follow-up with CMR-Late Gadolinium Enhancement (LGE, panels on the left), fused PET/CT images (middle panels) and PET images with delineated myocardial uptake volume (MUV, red contours, panels on the right), in two patients without significant PET inflammation at 4 months (A and B, FU.PET-) and four patients with PET inflammation at 4 months (FU.PET+, panels C to F). Two of the FU.PET+ patients with expansion of MUV from baseline-to-4-months (expanding FU.PET+, panels C and D) and two patients with no MUV expansion (nonexpanding FU.PET+, panels E and F). Individual LV.EF percentages and MUVs observed are indicated in white and red text, respectively. The upper levels of the gray and color scales for PET images are set to 3x the blood SUVmax.

Among all clinical and biological variables, only the 4-month high-sensitivity troponin-Ic plasma level exhibited a significant difference between the 5 expanding FU.PET+ patients and the other patients (13.0 [7.5-22.00] ng/l vs. 3.0 [2.0-9.5] ng/l, p=0.045) (see graphical abstract). It should, however, be noted that the expanding FU.PET+ patients also tended to be older (31.9 [27.6-42.5] years vs. 24.7 [21.7-31.9] years, p=0.055) and had higher body-mass-indexes (31.9 [27.6-42.5] kg/m^2^ vs. 24.9 [22.5-28.5] kg/m^2^, p=0.055).

## Discussion

We recently identified that a myocardial somatostatin-PET uptake volume criterion (i.e. > 18 cm^3^ of LV volume above 150% of blood SUVmax) was highly specific (94%) and sufficiently sensitive (84%) to detect acute phase myocarditis (*4*). The current study expands on this finding and shows that this volumetric PET criterion persisted 4 months after an apparently uncomplicated AM in as many as 41% of patients (FU.PET+ group). This rate is dramatically higher compared to inflammation detected by CMR (increased T2 level) in the months following AM (*13*) and at the 4-month follow-up of the current study population (7%). This may raise questions over the presence of true inflammation at the 4-month mark in these FU.PET+ patients. However, our current study indicates that the FU.PET+ status is associated with a poorer recovery of cardiac function, thus providing supportive evidence of the pathological significance of this status. Although the EFs for both ventricles were similar between FU.PET+ and FU.PET-patients at baseline, their 4-month levels were clearly lower in the FU.PET+ group due to the lack of significant baseline-to-4 month improvement (see Table 2 and Figure 1). Furthermore, the recovery of cardiac function was even worse and associated with a higher 4-month troponin Ic concentration in a subgroup of 5 FU.PET+ patients for whom the volume of inflammatory cell infiltrate (MUV) was still expanding between baseline and the 4-month follow-up.

Our hypothesis is that this FU.PET+ status corresponds to a local form of cardiac inflammation (i.e. with no systemic inflammation which may increase CRP levels) and to sub-acute or chronic disease (i.e. with a normal T2, the CMR parameter may not be sufficiently sensitive for detecting chronic myocarditis (*1*)).

Our results also suggest that somatostatin-PET monitoring may differentiate patients that fulfill the AM PET criteria at 4 months (i.e. the FU.PET+ patients) due to prolonged active disease (i.e. the subgroup with expanding MUV and the highest troponin-Ic at the 4-month follow-up) from patients with a longer recovery time as a result of a more severe initial insult (i.e. the nonexpanding FU.PET+ patient group showing the lowest LV.EF and highest MUV at baseline (Figure 2)) (see graphical abstract).

The levels of high-sensitivity troponin-Ic achieved at the 4-month follow-up in the expanding FU.PET+ group are admittedly rather low, with a median of only 13 ng/l. However, concentrations of this order of magnitude are known to impact prognosis in the general population (*14*).

Our data and hypotheses are supported by the current understanding of somatostatin receptor expression in inflammatory cells. The somatostatin type 2 receptor (SSTR2), that [^68^Ga]Ga-DOTA-TOC binds to, are highly expressed in the two main cell types involved in myocarditis pathogenesis: activated T-lymphocytes (*15*) and monocyte-derived (also called CCR2+) macrophages (*16*). Activation of the T cell system is indeed believed to be the major pathophysiological mechanism underlying autoimmune myocarditis and autoimmune inflammatory cardiomyopathy (*1,17*). Further downstream in this pathway, the CCR2+ macrophages mediate the greatest inflammatory effects in myocarditis and contribute more generally to myocardial inflammation and heart failure pathogenesis. These types of macrophages have previously been found to infiltrate the hearts of patients suffering from Cocksakievirus B infection-related myocarditis (*18*) and from severe myocarditis induced by immune checkpoint inhibitors (*19*). CCR2+ macrophages are also associated with persistent LV systolic dysfunction and adverse LV remodeling following mechanical unloading in heart failure patients (*20*). Finally, these macrophages may also play a critical role in heart failure progression after myocardial infarction particularly in non-infarct regions (*21*). These observations support that macrophages constitute a major therapeutic target for cardiomyopathy caused by myocarditis but also of other origins (*22*).

Only three of our patients had an LV.EF of < 50% at follow-up. Further studies on more seriously ill populations are therefore warranted to determine whether somatostatin-PET imaging of lymphocytes and macrophages may be an alternative to myocardial biopsy for detecting inflammatory cardiomyopathy (*2*). This information is essential before prescribing any immunosuppressive treatments (*2*).

Our results also raise questions about current recommendations which limit physical activity and sport for 3 to 6 months in patients with normal follow-up results and in patients with a limited scar pattern assessed by CMR at follow-up (*23*) since a number of such patients in our study showed signs of a significant inflammation volume by somatostatin-PET at the 4-month follow-up.

The small sample size of our study population and specifically of the two sub-groups of expanding and nonexpanding FU.PET+ patients constitute a major limitation. Results therefore need to be confirmed in larger populations.

## Conclusion

It is not uncommon for somatostatin-PET to detect myocardial inflammation at 4 months from an apparently uncomplicated AM. Our current study demonstrates that this affected 41% of patients. Myocardial inflammation detected by somatostatin-PET was associated with poorer recovery of cardiac function, suggestive of more persistent active disease or a longer recovery due to more severe initial insult. These new findings have the potential to improve monitoring and treatment adaptations for myocarditis patients.

## Acknowledgements

The authors thank Dr. Petra Neufing for critically reviewing the manuscript and the staff of Nancyclotep (https://nancyclotep.com) for technical support.

## Independent data access and analysis

Pierre-Yves Marie has full access to all the data obtained in the study and is responsible for its integrity and analysis.

## Data availability

The raw data that supports the findings is available from the corresponding author upon reasonable request.

